# It Takes Two to Tango: Supporting Public Health Leaders to Effectively Engage Political Leadership During Crises

**DOI:** 10.64898/2026.07.27.26350454

**Authors:** Marine Buissonnière, Ethan Guillén, Amanda McClelland, Kayla Jashinsky, Jobin Abraham

## Abstract

Nineteen individuals with diverse public health leadership experience across different levels and domains, from countries with varying levels of economic development, and who have direct involvement in crises were interviewed. A concise literature review was conducted, examining the intersection of politics and public health leadership, as well as a brief review of relevant existing leadership courses, training programs, and fellowships. The interviews were transcribed in full, anonymized, coded (total 1104 coded excerpts, 762 discrete excerpts), and used as the basis for analyzing findings.

Our research identified four thematic areas at the intersection of public health and political leadership in public health crises: attributes and skills; role of and threshold for political engagement; structural elements underpinning effective public health response; and modalities for building public health leadership. Public health leaders need to learn core skills, especially effective communication, diplomacy, and advocacy, and cultivate key attributes, especially integrity and humility, that support their ability to engage with political decision makers. Political leaders need to contribute productively ways, and at the appropriate time, to ensure societal cohesion and facilitate a well-resourced multisectoral response with public health at its core. Pre-established relationships and coordination mechanisms can play a supporting and productive role when crises strike. Strategies that incorporate experiential learning, peer collaboration, and mentorship are central to building the skills public health leaders need to succeed in crises.

Many tangible steps can be taken to foster and support public health leadership including: integrating the political dimensions of public health into public health education; tailoring public health trainings to include practical skills such as stakeholder engagement, effective communication, negotiation, policy advocacy, and community engagement; creating supportive ecosystems through peer, mentor, and fellowship approaches; and advocating for strengthening the structural interface between politics and public health.

## INTRODUCTION

Throughout the COVID-19 pandemic, countries with varying levels of resources and reported preparedness struggled to adequately respond. This was often attributed to failures of political leadership that negatively impacted public health system performance. Poor political leadership can render even sound public health recommendations ineffective when evidence is disregarded or neglected, divisive messaging is communicated, and public health is used as a scapegoat. Worse, poor political leadership can amplify poor public health policy and practice.

However, there are many examples of effective responses, where public health leaders effectively engaged and leveraged political decision-makers by working with them to involve communities and maintain social cohesion, rally impacted sectors, and make decisions based on evidence and context, ultimately leading to better public health and societal outcomes. Given that politics inevitably permeates public health crises, it is critical that public health professionals learn how to effectively navigate such situations. In addition to technical skills, public health leadership also needs to gain specific “soft” skills and learn from the experiences of others so that they can more effectively tackle critical non-public health tasks they will face during crises.

Our research is intended to identify the skills and attributes, as well as the structural environments, that form the basis for effective engagement between politicians and public health professionals. It also seeks to identify the extent to which those elements map onto existing training or capacity building programs offered to public health professionals, and how they could help inform and refine existing or new initiatives to strengthen the public health workforce. Finally, we aim to point to structures likely to enable successful engagement between public health and political leadership, as well as strategies public health leaders can apply for successful advocacy or negotiation to establish these structures.

## MATERIALS AND METHODS

### Data collection and analysis

We conducted semi-structured interviews to gain insights into leadership considerations involved in successfully navigating politics and public health during crises. Nineteen interviews were conducted, primarily via video call with some in person, over a three-month period (September-November 2023). An interview protocol standardized the data collection process, with open-ended questions used to elicit detailed responses.

Thematic analysis techniques were employed to analyze the qualitative data collected. Interviews were transcribed in full, anonymized, and coded based on initially anticipated themes. As interviews continued, themes were adjusted according to emerging patterns and perspectives. A final coding scheme includes 4 themes and 37 sub-themes. Coded interview excerpts (1104 total coded excerpts, 762 discrete excerpts) were used to analyze findings. Researchers identified, discussed, and solidified patterns, connections, and variations within and across themes.

### Selection of interview subjects

We approached 25 public health and political leaders via email, based on their diverse and extensive leadership experience across different levels and public health domains, and in countries with different levels of economic development. We established our list of potential interviewees based on past professional interactions, their body of peer-reviewed literature and published reports, and media references confirming their direct involvement and relevant experience at the intersection of public health and politics during public health crises. Of these, 19 individuals made themselves available for interviews.

These 19 leaders met one or more of these established criteria: currently hold/have held senior positions as civil servants or political appointees in health ministries, including at ministerial levels, or have served as health ambassadors; direct/have directed national public health institutes (NPHIs) or acted as International Health Regulations (IHR) national focal points, or both; have been designated as incident managers for responding to public health crises; hold/previously held public health leadership positions at national, regional, district, or city levels; or lead/played significant roles within the United Nations (UN), international or regional organizations, international nongovernmental organizations, or global philanthropies.

Participants collectively bring experience working at regional and global levels, and in-depth experience in various countries in Africa (West Africa: Ivory Coast, Liberia, Nigeria, Senegal, Togo; Central Africa: Cameroon, Democratic Republic of Congo; East Africa: Ethiopia, Kenya, Uganda); North America (Canada, United States); Europe (France); and South Asia (Pakistan). Collectively, they possess extensive experience and were in leadership positions during various public health crises over the past decade, including the COVID-19 pandemic. Out of the 19 interviewed individuals, all trained as clinicians; 15 also possess advanced professional degrees. There were 11 male and 8 female interviewees.

### Literature and course review

We conducted a literature review examining the intersection of politics and public health leadership to develop an evidence-based understanding of factors essential for effective political and public health interactions during public health crises, which provided a foundation for the study and directed formulation of interview questions. We also conducted a review of existing leadership courses, training programs, and fellowships tailored for public health professionals to gain insight into available educational and learning opportunities, with a focus on identifying potential gaps in the leadership needs of public health professionals.

### Ethical considerations

The Resolve to Save Lives research committee reviewed the research design and determined the research to be exempt from the requirements of 45 CFR 46 under the following category: Exemption 2 – 45 CFR 46.104(d)(2) – Research that only includes interactions involving educational tests (cognitive, diagnostic, aptitude, achievement), survey procedures, interview procedures, or observation of public behavior (including visual or auditory recording) meeting the following criterion: (ii) Any disclosure of the human subjects’ responses outside the research would not reasonably place the subjects at risk of criminal or civil liability or be damaging to the subjects’ financial standing, employability, educational advancement, or reputation.

## RESULTS

Our research identified four thematic areas at the intersection of public health and political leadership in the context of managing public health crises: 1) public health leadership attributes and skills; 2) the role of and threshold for political engagement during crises; 3) structural elements underpinning effective public health response; and 4) modalities for building public health leadership. Under each thematic area, sub-themes were derived based on how frequently interviewees cited them (Table 1).

**Table 1.** Themes and subthemes mentioned in interviews.

| **Theme** | **Subtheme** |  | **Mentions** |
| --- | --- | --- | --- |
| **Public Health Leadership Attributes & Skills** | Attributes | *Integrity* | 57 |
|  |  | *Humility* | 32 |
|  |  | *Empathy* | 17 |
|  |  | *Composure* | 9 |
|  |  | *Curiosity* | 4 |
|  | Skills | *Effective communication* | 102 |
|  |  | *Diplomacy and advocacy* | 55 |
|  |  | *Community concern* | 43 |
|  |  | *Decisiveness and proactive action* | 33 |
|  |  | *Collaboration and team building* | 27 |
|  |  | *Analytical abilities* | 24 |
|  |  | *Active listening and inquiry* | 12 |
|  |  | *Systems thinking* | 12 |
|  |  | *Technical competency* | 11 |
|  |  | *Being goal oriented* | 9 |
| **Role of and Threshold for Political Engagement in Public Health Crises** | Political leadership | *Communicating with the public and all sectors* | 45 |
|  |  | *Decision-making on matters beyond public health, support for decision made by public health leaders* | 33 |
|  |  | *Resource allocation and mobilization* | 27 |
|  |  | *Elevation of issues* | 25 |
|  |  | *Accountability and transparency* | 13 |
|  |  | *Multisectoral coordination* | 11 |
|  |  | *Activating appropriate legal powers* | 7 |
|  |  | *Fostering collaborative partnerships* | 5 |
|  | Engagement Threshold | *Fear and outrage* | 16 |
|  |  | *Size and impact of crisis* | 10 |

| **Theme** | **Subtheme** | **Mentions** |
| --- | --- | --- |
| **Structural elements underpinning successful political and public health alignment** | Political-public health interface | 107 |
|  | Multisectoral coordination | 51 |
|  | Community relations | 40 |
|  | Coordination across levels of government | 36 |
|  | Preparedness | 26 |
|  | Clarity of roles and responsibilities | 11 |
|  | Effective managing of partnerships | 11 |
| **Modalities for building public health leadership** | Hands-on experience | 60 |
|  | Leadership training | 50 |
|  | Academic educational opportunities | 26 |
|  | Peer-to-peer support | 22 |
|  | Mentoring, coaching, and fellowships | 17 |
|  | Professional affiliations | 8 |

These four themes, taken together, encompass core elements that enable successful management of public health crises. Public health leaders need to learn appropriate technical and “soft” skills and cultivate attributes that support effective leadership and political engagement. Political leaders need to contribute productively and support a well-resourced whole-of-society response with public health at its core. Underpinning these individual roles and traits are structural elements that support working together both day-to-day and during crises. Building public health leadership highlights modalities for supporting development of effective leadership skills and attributes.

### Public health leadership attributes and skills

Certain attributes of individual public health practitioners become critical for effective leadership in times of crisis to ensure effective alignment between politics and public health. In our interviews, integrity, humility, empathy, composure, and curiosity emerged as indispensable traits (in order of number of mentions). Core trainable skills considered to contribute to public health leadership success included: effective communication; diplomacy and advocacy; community concern; decisiveness and proactive action; collaboration and team building; analytical abilities; active listening; system thinking; technical competency; and being goal oriented.

### Role of and threshold for political engagement

Political engagement in public health crises is necessary as it is unrealistic for public health to be considered separately from politics. Primary political roles include: communicating with the public and societal sectors; supporting decisions made by public health leaders including those beyond the purview of public health; allocating and mobilizing resources for response; appropriately elevating issues; ensuring accountability and transparency; facilitating multisectoral coordination; activating appropriate legal powers; and fostering collaborative partnerships. Although political authorities should generally be engaged at some level regardless of the level of crisis, thresholds for engagement should shield routine public health work from political interference, which should factor in the amount of public fear or outrage the crisis may generate and the size and impact of the crisis on the public health system and society.

### Structural elements underpinning successful political and public health alignment

While political leadership hinges in part on the capabilities and actions of individuals, several structural elements are needed for successful public health response: interface between politics and public health; multisectoral coordination; community relations; coordination across national, regional, and local levels of government; preparedness; clarity of roles and responsibilities; and effective managing of partnerships.

### Modalities for building public health leadership

Key attributes for building public health leadership include: hands-on experience; specialized leadership training; academic educational opportunities; peer-to-peer support; mentoring, coaching, and fellowships; and professional affiliations.

## DISCUSSION

### Leadership attributes and skills

Several themes emerged around leadership qualities of successful public health and political leaders that are considered essential for effective public health response, with a particular focus on those needed to optimize interactions between public health and politics during a crisis. These comprise attributes (forming the basis of a leader’s character that contribute to effectiveness) and skills (abilities learned and developed through training, education, and experience). Structural elements, including various systems and processes, were are also factors that may facilitate or impede demonstration of leadership attributes and implementation of leadership skills in crisis.

Although we discuss attributes and skills separately, they are interconnected and work in tandem. Leadership attributes form the foundational character of a leader, while skills allow for consistent and appropriate execution. Research suggests that skills can be built,^1,2^ and attributes cultivated through effort.^3,4^ As an illustration, humility encompasses recognizing self-limitations, acknowledging the contributions of others, and maintaining a receptive attitude toward feedback, all of which can be improved through self-reflection and enhanced through targeted training.

#### Leadership attributes

Integrity, humility, empathy, composure, and curiosity emerged as indispensable traits of successful leaders, each playing a unique role in navigating crisis-related challenges, including the relation between politics and public health.

##### Integrity

The attributes of trust and credibility reinforce each other and are important to steer impactful responses. Effective leadership correlates with integrity, increasing effectiveness.^5^ Truthful language, impactful actions, and consistent conduct are key factors in establishing a sense of reliability and authenticity, ultimately building to credibility that instills trust in political decision makers, leaders from other sectors, and communities, and fosters support for and adhesion to public health recommendations.

##### Humility

A widely recognized attribute in public leadership,^6^ humility signals a demand for leaders who are self-aware, recognize diverse skills and perspectives, are unafraid of admitting mistakes and making necessary pivots, approach their roles with minimal ego, and focus on service rather than self-promotion.

##### Empathy

Leaders who can understand and connect with the experiences and emotions of others (including political opponents) and of communities foster inclusive culture and proactive ways to address divisions.^7^ Medical training and frontline clinical experiences can contribute to compassionate leadership style. Gender may influence a leader’s capacity to lead with empathy, consistent with findings that show women leaders outperformed men in key areas such as learning agility, communication, and collaboration,^8^ and those of a separate study that found women more effectively managed communications than their male counterparts.^9^

##### Composure

Public health leaders who maintain a calm and collected demeanor in challenging situations (noted in prior health crises including SARS^10^) can instill confidence in affected communities and society.

##### Curiosity

Leaders should display a genuine interest in learning and adapting as situations evolve.

#### Leadership skills

Public health leaders should be able to communicate effectively, navigate complex political environments, embrace the centrality of communities, act decisively in uncertain conditions, foster collaboration, actively listen, use analytical thinking, consider systemic implications, and maintain a goal-oriented mindset.

##### Effective communication

This essential skill for public health leaders is a major weakness in many. (Table 2 provides examples of poor and best-practice communication tactics.) Public health leaders need to interact with elected and appointed officials and decision-makers in diverse sectors whose support they need to respond to public health crises, and with the broader public with whom they must maintain a relationship based on trust.

**Table 2.** Poor and best practices that relate to effective communication.

| Poor practices | Best Practices |
| --- | --- |
| • Failing to translate scientific concepts into clear and understandable language for a lay audience.<br>• Assuming that merely simplifying the language will suffice to ensure universal understanding.<br>• Overloading people with excessive information; delivering lectures.<br>• Using paternalistic approaches and failing to address audiences as adults.<br>• Defaulting to ill-timed, linear, and one-dimensional approaches, dominated by a single (and at times inappropriate) messenger.<br>• Communicating in a language unfamiliar to the audience.<br>• Failing to strike a balance between overly cautious language (common in public health) and unwarranted certainty in the face of evolving evidence.<br>• Failing to acknowledge that effective communication is not a standalone solution. Without tangible efforts to solve problems, even the most eloquent communication is ineffective. | • Being an interpreter of science, which involves the translation of scientific information into clear messages for elected officials and the public.<br>• Knowing how to outline the political, social, and economic ramifications of a public health crisis to politicians. Leading with selected evidence and showing what happens if what you are asking for materializes, and if it does not.<br>• Crafting straightforward messages, while leaving room for needed adjustments over time without creating contradictions.<br>• In the face of evolving evidence, using a communication approach which is transparent about what is known (and how), what is unknown, what would be desirable to know, and when we might know.<br>• Using credible frames and persuasive language adjusted for the audience, delivered by an appropriate messenger.<br>• Limiting the number of messages in a speech or a response, the length of speaking time on TV, and the length of written materials when targeting high-level individuals to provide a top-line summary of key messages.<br>• Leveraging the role and power of storytelling to build a connection with the audience.<br>• Developing multi-faceted communication strategies which include multiple channels (including local and social media) and community-level engagement through widely-used communication channels and respected leaders, including women, youth, and faith-based leadership.<br>• Being prepared, patient, persistent, and repetitive. |

Most public health professionals do not receive comprehensive communication and media training earlier in their careers, leaving them ill-prepared for the communication requirements of leadership positions. Communication trainings tailored to public health practitioners should cover effective data presentation, including concise explanation of key information and clear actions to be taken. Focusing on message content and delivery is essential, but insufficient; programs should also cover key strategies to understand and relate to audience circumstances and values, including appealing to emotions through relatable storytelling. Regular opportunities to interact with various and potentially indifferent or hostile audiences should be sought out.

##### Diplomacy and advocacy

Public health leaders must adeptly navigate complex situations and champion public health recommendations with diverse political stakeholders and decision-makers, during and in between crises. Effective communication is a component of diplomacy and advocacy, and include a spectrum of strategic activities aimed at influencing political decision-makers, evolving policies, securing resources, and driving tangible change. They typically require a blend of analyzing political landscapes and power dynamics, mapping relevant stakeholders, carrying out strategic engagement, and communicating proficiently. (Table 3 provides examples of poor and best practices related to diplomacy and advocacy.)

**Table 3.** Poor and best practices that relate to diplomacy and advocacy.

| Poor Practices | Best Practices |
| --- | --- |
| • Lacking an understanding of the context and how decisions are made, when, and by whom. Not grasping who you are engaging with.<br>• Failing to understand the sources and distribution of power, who the true decision-makers are, who/what they are influenced by, and how to sway them so that they act or move aside.<br>• Being uninterested in politics, politicians' interests and values, and incentives underpinning political decision making.<br>• Naively assuming that if something makes sense, it will automatically happen, without rigorously considering the intricate intersection of politics and public health.<br>• Remaining confined to a technical public health language, which generally does not resonate with politicians.<br>• Merely presenting politicians with facts (and more facts) and recommendations without engaging in their world. Not translating public health findings into compelling points for politicians.<br>• Failing to disseminate public health research outcomes beyond the public health world.<br>• Believing that negotiating is outside the purview of public health professionals and unrelated to their field. | • Actively listening to others. Gathering as much information as possible from as many people as possible.<br>• Developing an understanding of the system, including to whom and at which time it is the most appropriate to reach out, and understanding the correct sequencing for maximal impact (who to engage first, etc.).<br>• Understanding the motivations of politicians: What drives them? What do they care about? How do they wish to be perceived? What would benefit them, their communities, those who (re)elected them? Leading with the issues they are concerned about.<br>• Never underestimating the person to whom you speak.<br>• Based on who you are speaking with, adapting your messaging to place your priorities in a context they care about. Adjusting your language, the delivery of your message, and the amount and the sequence of information provided.<br>• Creating a tipping point to compel politicians to act, through highlighting the gains if they do, or the risks if they don't, and understanding how to leverage public opinion.<br>• Being aware of the impact of what you are saying/doing, understanding who you may be upsetting, gauging how far you can push, and mitigating potential consequences.<br>• Learning tactics, including going on a trip with a leader (where they may be more accessible) and getting to know someone on their senior team. |

Although public health is often politically contextualized and constrained, diplomacy and advocacy skills are by and large absent from public health education curricula. Public health leaders often receive no training in this area, instead learning these skills primarily through professional experience. Such programs would have most impact if they used integrated learning approaches that include theory as well as case studies and practice.^11^

##### Community concern

Political and public health leaders must be attuned to realities and needs of communities to generate the engagement needed for successful public health responses. For example, how well countries performed on COVID-19 was not about pandemic preparedness, democracy, income inequality, universal health care, or hospital capacity, but people’s trust in each other (and government), underscoring the inherent communal dimension of any successful public health response.^12^ (Table 4 provides examples of poor and best practices related to community concerns.) There was greater resistance to attempts to sideline or marginalize public health guidance where public health had preestablished trust from communities.

**Table 4.** Poor and best practices that relate to community concern.

| Poor practices | Best Practices |
| --- | --- |
| • Remaining too lofty, failing to purposefully engage at community levels because of the complexity of reaching depth and scale.<br>• Not sufficiently investing in the depth and texture of local areas. Dismissing local beliefs and perceptions.<br>• Ignoring communities in normal times and only rushing to build relationships in crises.<br>• Failing to adequately engage with communities and their diverse set of leaders.<br>• Providing insufficient explanations and expecting communities to simply trust public health workers and guidance.<br>• Failing to recognize that public health requires behavior change, which does not happen instantaneously or through edicts.<br>• Undermining community trust by failing to be transparent, withholding or manipulating data, or failing to dissipate concerns related to data manipulation, political gain, or other improprieties.<br>• Struggling to be perceived as evidence driven. Being seen as politically affiliated, especially when working in areas hostile to the government.<br>• Using unidirectional communication, without meaningful feedback or interaction.<br>• Using buzz words and language that is not the vernacular of the community.<br>• Constantly soliciting information from communities about their needs without taking effectively addressing them. | • Building relations with communities and their leaders, which requires spending time in communities to develop community adhesion.<br>• Mapping of communities, especially those marginalized/isolated from the mainstream, identifying leaders, information sources, etc.<br>• Establishing relationships with communities and their leaders in ordinary, non-emergency times. Pre-existing relationships become especially important in crises, especially when restrictive measures are considered.<br>• Doing the research to build cultural awareness of local areas, making an effort to understand the sensitivities, local culture, and traditions. Investing in surveys and socio-anthropological research to understand: What do people think? Why might they have different attitudes? What enables them to believe what they believe?<br>• Taking the time to explain (with the right frame and language) public health perspectives and needed interventions; being truthful and transparent, human, and compassionate.<br>• Working with the right messengers. Scientists and community leaders may be more effective than politicians due to pre-existing credibility.<br>• Embracing community engagement as a two-way process between equals. Listening to views and concerns and co-constructing what feasible interventions look like.<br>• Approaching community engagement as a multifaceted non-linear exercise. Community radio stations and other key media, community leaders, women’s associations, and youth groups all need to be made stakeholders so that everybody feels involved and sufficiently empowered to have a say in problem solving.<br>• Ensuring that some measures are taken to address community needs. |

##### Technical and analytical skills, decisiveness

Leaders must be able to analyze vast amount of evolving information to support evidence-based recommendations, which proved especially challenging during COVID-19. Public health leaders who master this technical function instill confidence among politicians. Public health leaders should not only be capable of analyzing data, but also of acting decisively under time pressure and providing timely recommendations to political decision makers amid the potential for significant loss of life. When faced with uncertainty, the tendency is to delay rather than act quickly. Social, economic, and political realities will influence decision-making, and compromises are often necessary to secure even partial action rather than no action at all. (Table 5 provides examples of poor and best practices related to technical and analytical skills and decisiveness.)

**Table 5.**
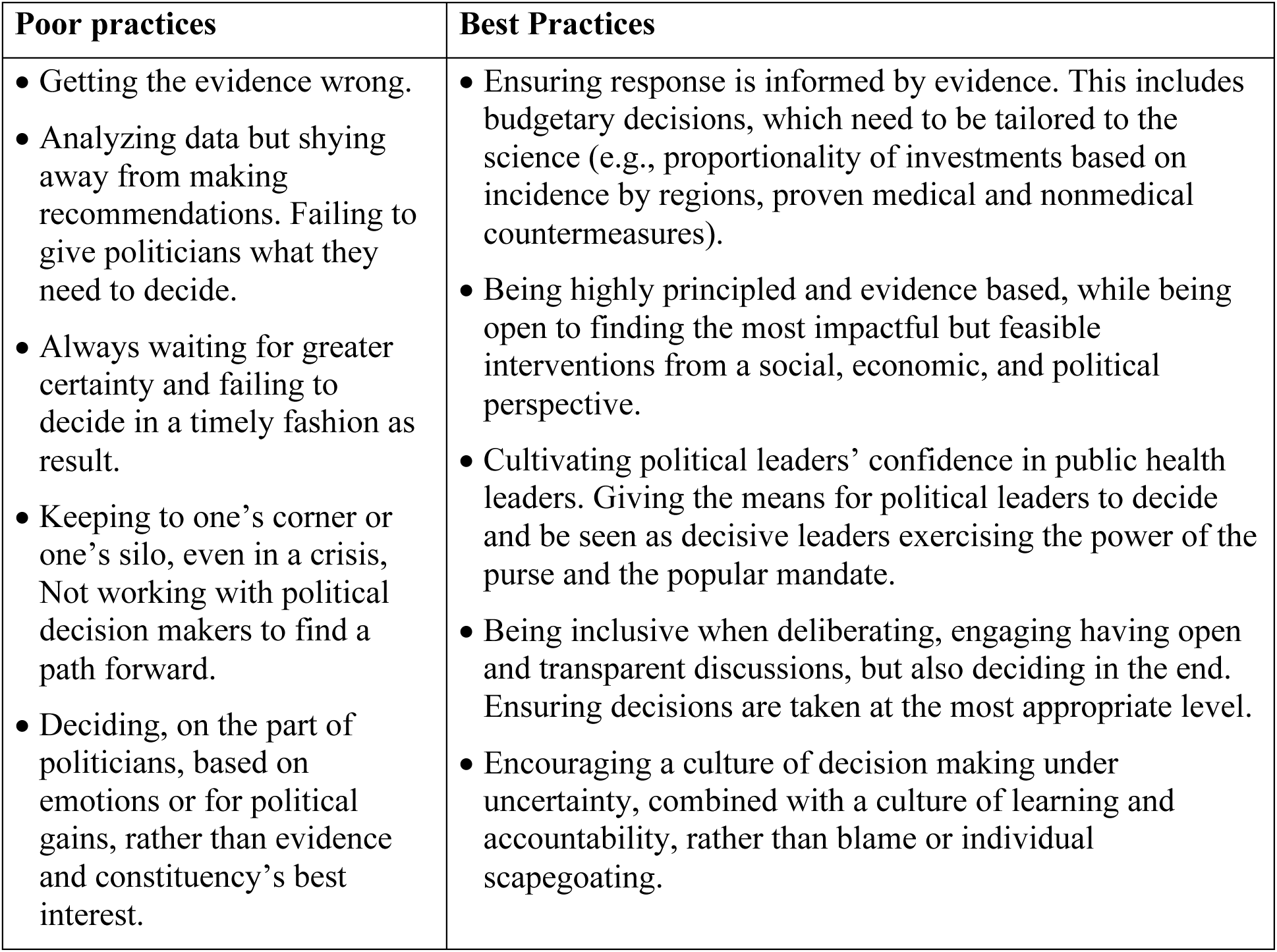
Poor and best practices that relate to technical and analytical skills and decisiveness.

| Poor practices | Best Practices |
| --- | --- |
| • Getting the evidence wrong.<br>• Analyzing data but shying away from making recommendations. Failing to give politicians what they need to decide.<br>• Always waiting for greater certainty and failing to decide in a timely fashion as result.<br>• Keeping to one's corner or one's silo, even in a crisis, Not working with political decision makers to find a path forward.<br>• Deciding, on the part of politicians, based on emotions or for political gains, rather than evidence and constituency's best interest. | • Ensuring response is informed by evidence. This includes budgetary decisions, which need to be tailored to the science (e.g., proportionality of investments based on incidence by regions, proven medical and nonmedical countermeasures).<br>• Being highly principled and evidence based, while being open to finding the most impactful but feasible interventions from a social, economic, and political perspective.<br>• Cultivating political leaders' confidence in public health leaders. Giving the means for political leaders to decide and be seen as decisive leaders exercising the power of the purse and the popular mandate.<br>• Being inclusive when deliberating, engaging having open and transparent discussions, but also deciding in the end. Ensuring decisions are taken at the most appropriate level.<br>• Encouraging a culture of decision making under uncertainty, combined with a culture of learning and accountability, rather than blame or individual scapegoating. |

##### Collaboration and team building

Public health challenges are inherently multifaceted, often extending beyond the purview of traditional health domains, requiring teamwork and cultivating collaborations across sectors and domains and at different government levels. (Table 6 provides examples of poor and best practices related collaboration and team building.) A public health leader adept at collaboration recognizes the interdependence of health with various societal sectors and understands their potential to bolster and amplify the effectiveness of the public health response. By leveraging the strengths of different sectors, a public health bridge builder can enhance the effectiveness of public health initiatives but also unlock innovative solutions, ensuring a more comprehensive and interconnected response.

**Table 6.** Poor and best practices that relate to collaboration and team building.

| Poor practices | Best Practices |
| --- | --- |
| • Oversized and unchecked egos, which destroy consensus and undermine people's goodwill to collaborate.<br>• Putting oneself forward, seeking personal gains, flag planting.<br>• Standing on the sidelines, waiting for others to fail.<br>• Seeing political and cross-sectoral engagement as a chore, even as it is a fundamental part of the job.<br>• Failing to understand and translate the impact (real and potential) of public health crisis on other sectors in terms other sectors can relate to and act on.<br>• Failing to effectively communicate across sectors and make allies of other sectoral leaders. | • Starting off with humility and recognizing one cannot do everything without collaboration.<br>• Building a cohesive team and maintaining a good team dynamic. Involving everyone in the collective effort.<br>• As a leader, properly delegating, recognizing the strengths of team members. Facilitating smooth operations to facilitate everyone being able to do their work.<br>• Recognizing that when holding a public health leader position, a large part of the job is political engagement and management.<br>• Building bridges across administrative levels, sectors, political divides. Being open to reaching out to people and talking with them. |

##### Other skills of importance

Active listening and inquiry encompasses hearing from many people from diverse backgrounds and paying attention to every argument, especially countervailing ones. System thinking involves absorbing information deeply and effectively, considering the broader context and implications of decisions, and collaboration across sectors. Goal orientation emphasizes the need to keep a clear focus on achieving objectives.

### Role of and threshold for political engagement

#### Political roles

Because public health cannot be entirely separated from politics, efforts should be aimed at optimizing the relationship between the two spheres, minimizing unnecessary interference, and building the competencies needed for public health practitioners to engage effectively in the political space. The political role in public health crises is multifaceted, and appropriate engagement can enable, or restrict, an effective public health emergency response.^13^

##### Communication

Political leaders must be able to: convey evidence-based guidance in the face of evolving information; address population concerns in a timely and forthright fashion; and preserve society’s cohesion during times of crises. Politicians who listen to public health practitioners are better able to understand technical or scientific guidance and convey it to the public. Political leaders who are good communicators tend to engage in two-way communication with communities and civil society, repeatedly traveling to affected areas and setting up channels for exchanges with the public, in contrast with one-way, top-down communication that is often the norm.

Communication by political leaders in times of crisis can bring people together by fostering a sense of security and safety, but can easily and quickly become counterproductive.^14^ The spotlight can be irresistible for political leaders, but the savviest ones understand when they are the right messenger and when others, including community or technical public health leaders, may be seen as more trustworthy messengers.^15^ Much depends on whether public health leaders find discerning, credible political leaders to partner with (e.g., New Zealand’s oft-cited COVID-19 response). When such partnerships are not possible, or when the perceived independence of public health advice may be undermined, public health leaders may need to keep their distance from elected leaders and even maintain their own communication channels with the public.

##### Decision-making

Effective public health measures do not exist in isolation due to societal and political constraints on implementation. Political figures need to back public health leaders’ recommendations, or constrain them as necessary, and make decisions beyond health across impacted sectors. While evidence and public health recommendations are key and politicians must not make decisions in a vacuum, public health leaders do not always have a full vision of the implications of various decisions that must be left to political leaders. Public health leaders need to think at the system level, while skilled politicians must factor in relevant parameters and gauge which measures will be acceptable to the population while maximizing public health impact. Lack of training or experience often leaves technical experts and academicians unfamiliar with the compromises inherent in political practice.^16^

##### Mobilizing and allocating resources

Although an essential role for political leaders,^17^ heads of ministries of health and national public health institutes often lack leverage with ministries of finance and other budget decision-makers to mobilize resources needed to address crises. Rapid deployment funding can be critical during emergencies, with standardized procedures allowing for immediate disbursement of resources. Engagement with higher political levels can speed availability both national resources as well as funds from foreign donors and development banks. However, just as important is advocacy for increased funding for longer-term response.

##### Elevating issues

Political leaders are responsible for calling attention to emerging concerns across all of society and ensuring they receive appropriate consideration. Political leadership has long been recognized as key in elevating the visibility of public health crises, both in positive ways to provide solutions,^18^ but also in negative ways that create needless fear.^14^ Political leaders should ensure that issues are taken up by the appropriate decision-making forum, and can also elevate issues through their own actions. In many cases, including during COVID-19, crisis management can be streamlined through use of ad hoc bodies that bypass existing bureaucracies.

##### Multisectoral coordination and partner engagement

Political leaders playing a central role in coordinating crisis response efforts across diverse sectors and partners is critical to public health response.^19^ In a crisis, a political leader at the highest level can engage all political leaders, across parties and regions, and summon them to work together. The head of government also has the power to bring together all ministries responsible for areas impacted by the public health crisis and establish clear lines of communication and responsibility.

##### Accountability and transparency

Political leaders should ensure transparency in decision-making, use of resources, and accountability for actions, which can increase trust by forcing information sharing and honest status assessments.^20^ Elected political leaders also carry a level of accountability that unelected public health officials do not. Lack of transparency and accountability by politicians can severely hamper a response, particularly when questions of financial improprieties and corruption arise that sow distrust.

##### Legal dimensions

Legal instruments must be in place so that public health institutions have appropriate authorities to discharge their crisis-related functions. For new situations (e.g., quarantines), needed legal instruments may not exist, and may require new texts (e.g., laws, decrees, or ordinances) for approval by government leaders.

#### Threshold for Political Engagement

There are diverging views as to when or how politicians should be involved in crisis. Some situations are entirely within the purview of public health and don’t require the political lever. In others, politicians should be engaged on an ongoing basis to build relationships and trust. Pre-existing relationships are key to aligning these spheres during crises, and should be forged before a crisis strikes to preclude the need to bring political leaders up to speed as it unfolds.

##### Size and potential impact

Politicians should step in when a crisis reaches a scale that significantly affects the population, the health system, or other key societal sectors. Political engagement at the highest levels of government, which in some cases will be smaller subnational administrative or political units, may be needed to address crises with potentially widespread consequences

##### Fear and outrage

Politicians should be actively involved when public sentiment is heightened by emotions and concerns, even if the risk may not be large. Collective recognition of these thresholds in size, impact, and public sentiment underscores the nuanced decision-making process that determines when political involvement becomes imperative in navigating and mitigating the effects of a public health crisis.

### Structural elements

While political and public health leadership hinges in part on individual training and experience, several structural elements are critical to form an integral framework essential for these spheres to work collaboratively in crises to orchestrate a successful public health response.

#### Political-public health interface

Engagement and alignment of political and public health leadership is critical.^16^ This includes establishing relationships before a crisis, sustaining exchanges in normal times, and building up the public health savvy of political leaders. Institutionalized mechanisms with various levels of formality can enable exchanges over time and set solid grounds when crises strike, particularly as individuals in key roles often move to other positions of responsibility and are replaced.

Parallel ad hoc systems are often disconnected from or sideline existing public health leadership and should be avoided.^21^ However, the ability of standing systems to perform under pressure must be assessed. In the case of COVID-19, some structures set up prior to the pandemic were not prepared for a long and all-encompassing state of emergency. The use of a “czar,” a high-level official who is granted broad, overarching powers to address a particular issue, can be an alternative to relying on a high-level platform to ensure the liaison between heads of state and all needed stakeholders and streamline communication and decision-making.

#### Multisectoral coordination

Organizing efforts across diverse sectors and mechanisms is closely intertwined with the need for coordination at the strategic level. Multisectoral coordinating fora are key to response success, with many failures linked to the absence of a standing body that promotes coordination and communication amongst parties and societal sectors.^21^ During COVID-19, many countries recognized and embraced such multisectoral coordination; 98% included “country-level coordination, planning and monitoring” in preparedness and response plans.^22^

#### Community relations

Standing relationships with communities, including traditional community leaders and religious authorities, can be key in successful outbreak response.^21^ Creating fruitful, long-lasting relationships with communities is multifaceted and includes understanding the communities being entered into, speaking in culturally-aware ways, building trust, and recognizing resource constraints and providing support when necessary.^23^

#### Coordination across levels of government

Effective links between regional, national, and local efforts help facilitate smooth cooperation and limit duplication across levels of governance. Multilayered coordination can be complex and tension-ridden, requiring harmonization from the highest political levels. Finding the optimal locus of decision making, and related control of resources, is necessary but can be fraught with tension. Countries, especially those with a federal structure, may not have established systems for systematic and coherent information flows or unified adoption of common guidance or approaches. This requires balance between central direction and local ownership to enable elected officials closest to the problem – often at the least resourced administrative levels (e.g., health zones, municipalities) – to share responsibility. Regardless of how the system is organized, alignment, support, and buy-in across all levels of government, especially at local levels, is important.

#### Preparedness

Being prepared well in advance of a crisis, including establishment of coordinating bodies and financing mechanisms for rapid response, is crucial. All too often, preparedness is shortchanged after a public health emergency passes as political leaders lose interest (the “panic-neglect” cycle), indicating a need for advocacy skills for public health officials.

#### Managing partnerships

Effective collaboration with international stakeholders requires understanding as to which are partners and allies, which may have agendas at cross purposes to those of another country or other stakeholders, and the specific mechanisms each uses to advance their objectives. During COVID-19, while many partnerships supported country responses, the complexity of these partnerships was on full display, with many countries reporting deficiencies from international partners including in the amount and type of support.^21^

#### Clear roles and responsibilities

Collaborating partners should avoid waste of resources and duplication of efforts, and properly document all actions taken.^21^ While the emphasis on needed linkages between different government bodies and external partners in a response is clear, there is also a need for mutual recognition and boundaries, particularly regarding the roles of public health and political leadership, enabling both to work towards the same objective.

Working directly in and with communities with an understanding of their cultural contexts is key. Interactions and decision-making need to be truthful and transparent, and preparations need to be thorough to enable a rapid response within communities when an emergency arises.

Although 95% of countries included community engagement related to risk communication in COVID-19 preparedness and response plans, key elements including ongoing provision of standard, non-outbreak healthcare services to communities were often lacking.^22^

### Building public health leadership

#### Hands-on experience

True leadership is forged in the crucible of practical challenges and decision-making, starting from the grassroots level all the way up to national leadership positions. Public health leaders without practical on-the-ground experience connecting with political leaders and building bridges with all stakeholders may have good technical skills, but may lack the textured understanding of working and making decisions at the local level. Experiential learning is associated with a large and significant increase in learning outcomes relative to non-experiential activities.^24^ Practical experience can impart operational savvy, knowledge about what works in real-life situations, and the ability to solve unexpected problems.

Professional pathways tend to be country-dependent, so seasoned leaders should create opportunities for experience building, including delegating responsibility for some tasks to junior staff to prime them to become leaders themselves. Research indicates that identifying new opportunities to gain hands-on experience were second only to a need for formalized training.^25^

#### Leadership training

Leadership skill building is generally not covered in most standard public health training programs. While some Doctor of Public Health programs leverage competency models, including leadership, as a cornerstone of their curricula,^26^ Masters programs typically do not; only 55% of MPH programs in the United States offered courses on leadership and just one taught crisis leadership.^27^ In the absence of clear pathways for well-rounded practitioners, those skills most often get built after a practitioner has already reached a leadership position.

Structured learning programs designed to hone specific leadership competencies should cover communications and media training, crisis management, and negotiation and advocacy. Bringing together leaders from public health and other proximate sectors can help create a common language and frames of reference to ease future coordination. Simulation exercises (e.g., case studies, role playing, negotiation simulations) are effective training methods to develop public health leadership skills.^28^

#### Academic education

Formal academic programs successfully educate future medical and public health professionals, providing theoretical frameworks and technical proficiency. Although some leadership programs exist for seasoned practitioners, significant gaps remain. Many programs are, at least in part, based in and focused on high-income countries and lack a curriculum with the capacity to address the full range of needs for the numbers of public health leaders required globally.^27,29,30^

#### Peer-to-peer interactions

Exchanging ideas with and learning from peers, formally and informally, is in many ways more valuable than formal training programs. Peer-to-peer (P2P) interactions can transcend country boundaries to provide sounding boards and promote problem-solving, and can also involve sharing scarce resources and expediting time-sensitive collaboration. Team-based approaches benefit from of group learning,^31^ which facilitates transfer of knowledge into practice.^32^

#### Fellowships, mentoring, and coaching

Personalized guidance from mentors fosters wisdom transfer and professional growth, and can extend after formal workplace relationships end. Mentors ideally should exemplify leadership qualities and embody essential attributes and skills, rather than primarily on technical proficiency or title.^33^ While mentoring and coaching are a key element of successful public health leadership development,^25,34^ and some fellowship programs stand out for their thoughtful design including their selection of mentors, this approach appears to be underutilized. Country-led initiatives that identify rising leaders for mentorship by current leadership could bridge this gap.

### Limitations

While our research offers valuable insights into the perspectives of the 19 individuals interviewed, it is important to acknowledge certain limitations that may impact the generalizability and applicability of the results. While interviewees represent a broad set of perspectives and positions, the sample size may be insufficient to fully encompass the diverse range of opinions among political and public health leaders. The strong representation of participants from North America, Africa, and Europe may limit the generalizability of the findings to other world regions. The qualitative nature of the interviews may lead to subjective interpretation and bias, as responses are inherently influenced by personal experiences and perspectives. Caution should be exercised in extrapolating conclusions to areas not adequately represented in the study.

### Recommendations

Based on the peer-reviewed literature and insights from interviews, we offer recommendations that span four overarching areas: leadership education, skill development training, leadership development, and structural elements (Table 7). By implementing these recommendations, donors and policymakers can enhance the leadership skills of public health professionals, their ability to optimize public health and political interactions, and the effectiveness of public health crisis responses.

**Table 7.** Recommendations to enhance leadership skills of public health professionals.

| **Implementation area** | **Recommendations** |
| --- | --- |
| **Enhance public health leadership education** | • Support academic public health curricula to include analytical frameworks and content that sensitizes students to the political dimensions of public health from the outset of their education and initiates them in analyzing and navigating these dimensions.<br>• Expand academic education opportunities to include advanced degrees or certifications in public health leadership. |
| **Develop practical skills of public health Professionals** | • Conduct a detailed review of existing trainings available to public health practitioners to identify missing leadership components that should be included, and assess accessibility to LMIC-based non-English speakers.<br>• Expand or develop tailored training opportunities for public health practitioners, especially those working in NPHIs or in public health functions at MoHs.<br>• Focus on key practical skills needed to engage with political leaders and navigate the political landscape, including stakeholder engagement, effective communication, negotiation, policy advocacy, and community engagement.<br>• Embrace blended learning approaches that match the learning patterns of working professionals. Encourage and incentivize practitioners to develop these skills early and continue honing them throughout their careers.<br>• Involve experienced public health leaders from LMICs in training development and delivery, with a focus on localized offerings in outbreak-affected regions and subregions and in local languages. |
| **Support public health leaders' growth** | • Support and expand activities of peer-to-peer platforms and fellowship schemes that foster formal and informal exchanges, learning, mentoring, and relationship building among emerging leaders (i.e., those on the path to leadership positions) from regions or subregions and explore the need and value of creating additional opportunities.<br>• Optimize mentorship programs by finetuning selection of mentors to ensure that they offer the right combination of leadership attributes and skills, relevant public health background, and a commitment to coaching mentees (rather than relying on titles and technical skills alone).<br>• Explore and conceive of ways experienced leaders with relevant track records could be mobilized and incentivized to provide tailored mentoring to emerging public health leaders, particularly within MoHs and NPHIs. |

| Implementation area | Recommendations |
| --- | --- |
| **Strengthen the structural interface between politics and public health** | • Foster joint training and collaborative initiatives between political and public health leaders during non-crisis times to build relations, demystify each other's world, and create shared language and mental models around decision-making and communication.<br>• Adapt or create evidence-based negotiating and advocacy frameworks to support the effective engagement by public health practitioners in ways that reflect their values and motivations. This may also require academic research on the types of public health messaging that engages varying audiences (e.g., heads of government, Ministry of Finance, Ministry of Agriculture, etc.).<br>• Advocate for and support the establishment of scalable country and subnational level coordination mechanisms, rooted in public health expertise, for the appropriate level of political engagement, multisectoral coordination, and alignment of all governance layers.<br>• Conduct further research on the interplay of politics and public health in LMICs to expand the existing literature, which is largely focused on HICs. |

## CONCLUSION

The public health and political leaders interviewed identified key attributes and skills essential for enhancing the alignment of public health and political leadership, most notably integrity, effective communication, and diplomacy and advocacy. They outlined specific roles for political actors and crisis thresholds at which they should intervene to support a whole-of-society response with public health at its core. They also pointed to structural elements that link the political and public health spheres to support enlightened leadership and underpin a successful response, and offered insights on ways public health practitioners can develop the competencies required to effectively navigate the political-public health interface. Taken together, the interviewees have shown a path ahead, supported by the broader literature, for building a cadre of effective public health leaders who can address the many complex crises the world faces.

However, the best public health leaders and the most efficient structures cannot, on their own, overcome the challenges posed by ineffective or destructive political leadership, as illustrated by the experiences of some countries during COVID-19. Despite existing challenges, there are many tangible, low-resource steps, outlined in the recommendations, that can be taken to develop and support current and future public health leaders. Given the incredible strains put on public health systems and leaders during COVID-19, it is more important than ever for governments and funders to take action so we can be better prepared globally to prevent and respond to future public health crises.

## Supporting information

Interview Questions

## Author declarations

All authors have seen and approved the manuscript, which has not been previously published by a journal.

## Ethics statement

All relevant ethical guidelines have been followed. All necessary participant consent has been obtained and the appropriate institutional documentation archived. The Resolve to Save Lives research committee reviewed the research design and waived ethical approval for this work as it determined the research to be exempt from the requirements.

## Data availability statement

All data relevant to the study are included in the article. To safeguard the privacy of the participants and those involved in their services, we cannot make transcripts available.

