## Supplementary material for "It Takes Two to Tango: Supporting Public Health Leaders to Effectively Engage Political Leadership During Crises": Interview Questions

The questions are written to be asked both of public health and political leaders to provide their different experience and perspective to the same issues, and their different perspectives on each other. The questions are meant to focus on leadership aspects of an effective response to not overly rely on technical considerations.

We’d like to start the interview asking generally about what makes for an effective public health response, both in terms of leadership and structures to link the public health and political leadership realms. What do you see as the key elements of an effective public health response (PHR) in a situation of crises (outbreaks, other PH threats) from a political/public health leader?

- What role does the engagement of political authorities play in a successful PHR?
- Is there a threshold of size of emergency at which political engagement becomes critical (i.e. is political engagement critical with smaller outbreaks)?
- What political and public health structures help support an effective public health response?
- How do different levels (e.g. federal versus local) of political and public health structures best work together to mount an effective response?
- What leadership qualities in public health/political leaders are most critical for a successful PHR? And for ensuring engagement across the PH / pol divide?
- What leadership qualities have you seen in political/public health leaders who have led to alignment between PH and political leadership, commitments at the highest levels and successful responses?
- How have successful public health leaders who display such qualities acquired those? What background, training or experience for public health/political leaders do you believe is most important?

Now we’d like to focus on a PHR you were involved in. What key elements at the nexus of political engagement and public health led to the successful response to X which you helped lead?

- What leadership qualities do you possess that led to a successful PHR in this case?
- How did you make the case to ensure sustained engagement at the required political levels? What was key to creating the right alignment between PH and pol leaders?
- What skillset did it take for public health professionals to make their case and be able to mobilize appropriate response across sectors and government structures and bridge the science / political divide?
- What training (beyond technical training) or other skill building vehicles have been important for you in leading this successful PHR?
- What training (beyond technical training) or other skill building vehicles do you wish you had had in leading a successful PHR?
- What training (beyond technical training) and leadership qualities did other key political/public health leaders involved in the response have? How had they acquired those?
- What were the most avoidable mistakes, at the nexus of PH and political engagement, which were made in responding to X?
- What skills did it take to engage this whole-of-society response?

Now we’d like to shift focus to ineffective PHRs you’ve witnessed. What are the key drivers of ineffective PHRs you’ve witnessed? (NB: Here we are not asking about Trump or Bolsonaro type failures where misinformation and populism drove failure. We are focused on less extreme cases.)

- How key are political failures versus failures of public health structures?
- What are the failures of political leaders/public health leaders that have led to poor responses?
- What are the top three things you’d suggest to avoid these types of failures?

If you were going to set up an ideal system for successful PHR that branched across the local, regional, national, and global level, what are the key elements you’d put in place?

- How would you delineate duties between political and public health leaders?
- What systems would align political and public health leaders in taking action?
- What training (or other means) would public health leaders have to bridge the divide and engage both political leaders and the public to support a successful PHER?
